# Greater early weight loss after metabolic/bariatric surgery predicts greater loss up to 2 years but is also associated with weight regain in the longer term

**DOI:** 10.64898/2026.10.07.26364977

**Authors:** Kavinraj Pushparajah, Dale Handley, Ashish Lobo, Sumaya Almansoori, Suzanne I. Alsters, Sanjay Purkayastha, Harvinder S. Chahal, Mieke van Haelst, Robin G. Walters, Fotios Drenos, Alexandra I. F. Blakemore

**Affiliations:** Faculty of Medicine, Imperial College London, London, UK; Department of Psychology, School of Biological Sciences, University of Cambridge, Cambridge, UK; Social, Genetic, and Developmental Psychiatry Centre, Institute of Psychiatry, Psychology and Neuroscience, King’s College London, London, UK; Department of Biosciences, College of Health, Medicine and Life Sciences, Brunel University London, London, UK; Department of Metabolism, Digestion and Reproduction, Imperial College London, London, UK; College of Medicine, Mohammed Bin Rashid University of Medicine and Health Sciences, Dubai Healthcare City, Dubai, United Arab Emirates; Genome Center, Department of Forensic Science and Criminology, Dubai Police GHQ, Dubai, UAE; South West Thames Regional Genetic Service, St George’s University Hospital NHS Foundation Trust, London, UK; Imperial Weight Centre, St Mary’s Hospital, Imperial College Healthcare NHS Foundation Trust, London, UK; Section of Clinical Genetics, Department of Human Genetics, Amsterdam University Medical Centers, Amsterdam, The Netherlands; Nuffield Department of Population Health, University of Oxford, Oxford, UK; College of Medicine, Nursing and Health Science, University of Galway, Galway, Republic of Ireland

## Abstract

**Background:** Early postoperative weight loss may help predict later outcomes, but its reliability for longer-term weight loss and regain remains unclear. We assessed whether the percentage total weight loss (%TWL) 3 months after metabolic/bariatric surgery predicted later %TWL weight change/regain.

**Methods:** This retrospective observational cohort study included 216 patients from the Personalised Medicine for Morbid Obesity cohort who underwent sleeve gastrectomy or Roux-en-Y gastric bypass and had 3-month %TWL and at least one later outcome. Linear regression assessed associations between 3-month %TWL and later %TWL plus weight change/regain outcomes. Adjusted incremental R^2^ quantified the additional variance explained by 3-month %TWL. Secondary procedure-stratified, diabetes-stratified, and baseline BMI comparator analyses are reported in the Supplementary Material.

**Results:** Greater 3-month %TWL was associated with higher 1-year and 2-year %TWL in adjusted models (both p < 0.001) but not longer-term %TWL (p = 0.078). Incremental R^2^ was 38.23% for 1- year, 11.75% for 2-year %TWL and 2.25% for long-term %TWL. Greater 3-month %TWL was also associated with greater recurrent weight gain from 1 to 2 years and from 1 year to long-term follow-up. In secondary analyses, findings were broadly consistent across procedure and diabetes subgroups, while 3-month %TWL explained more additional variance in 1-year %TWL than baseline BMI

**Conclusion:** 3-month %TWL predicted 1-year and 2-year outcomes but not longer-term %TWL, supporting its value as an early marker of postoperative weight trajectory. Heterogeneity in recurrent weight stability may contribute to weaker long-term prediction and warrants further study to determine whether early weight-loss patterns can identify patients requiring closer long-term weight management support.

**Key points:**

- Three-month %TWL predicted 1-year and 2-year %TWL.
- Greater early %TWL was associated with greater recurrent weight gain.
- Three-month %TWL explained more additional variance in 1-year %TWL than baseline BMI.

## Introduction

Weight-loss outcomes after metabolic/bariatric surgery (MBS) vary between individuals, even within the same procedure type^[1,2]^. Although many patients achieve marked weight loss, others have a less favourable initial response or subsequently experience recurrent weight gain^[3]^. Identifying early markers of these trajectories could support earlier, more individualised postoperative follow-up and identify sub-groups requiring careful long-term management. Across weight loss interventions, considerable emphasis is often placed on the initial weight-loss response, whereas long-term stability at the new weight may be a more important indicator of durable health benefit^[4]^.

Early postoperative weight loss has previously been associated with later outcomes. Manning et al. reported that weight loss at 6 weeks, 3 months and 6 months and weight-loss velocity between early intervals predicted maximal weight loss after sleeve gastrectomy (SG) and Roux-en-Y Gastric bypass (RNYGB)^[5]^. However, that study primarily examined maximal and 2-year weight loss, leaving uncertainty over whether a single routine measure, such as 3-month percentage total weight loss (%TWL) predicts fixed outcomes at 1 year, 2 years, and beyond 3 years as well as subsequent weight change.

Procedure type influences postoperative weight trajectories because SG and RNYGB differ anatomically and physiologically^[6]^, while pre-existing type-2 diabetes is associated with smaller postoperative weight loss^[7]^. Baseline BMI may be an important determinant of early postoperative weight loss: although %TWL expresses weight loss relative to baseline weight and is considered less dependent on starting BMI than percentage excess weight loss^[8,9]^, early postoperative %TWL may still be partly influenced by baseline BMI.

Our primary aim was to determine whether 3-month %TWL predicted later %TWL and subsequent weight change in a multi-ethnic UK cohort after SG or RNYGB. Secondary analyses examined whether associations were consistent by procedure type and diabetes status, and whether baseline BMI provided comparable predictive information.

## Methods

### Study design, cohort and ethical approval

This was a retrospective observational cohort study using data from the Personalised Medicine for Morbid Obesity (PMMO) cohort. Participants were recruited under informed consent within the PMMO study. Ethical approval was given by the NRES committee London Riverside (REC reference 11/LO/0935), and the research was performed in accordance with the Declaration of Helsinki. It is registered on https://clinicaltrials.gov/study/NCT01365416 with the clinical trial number NCT01365416.

A total of 2556 unrelated adults seeking NHS care for severe obesity were recruited between 2011 and 2021 for the Personalised Medicine for Morbid Obesity (PMMO) study. Participants had a BMI ≥ 40 or a BMI ≥ 35 with significant comorbidities. Of these, 1469 were recruited prospectively across different hospital sites. The present analysis was restricted to participants who underwent SG or RNYGB and had 3-month %TWL and at least one later outcome: 1-year, 2-year or long-term %TWL.

The final cohort included 216 patients: 113 underwent RNYGB, 103 underwent SG; 75 had diabetes and 141 did not. Outcome availability was 175 patients at 1 year, 126 at 2 years and 124 at long-term follow-up. Analyses used complete cases for each outcome. Thus, sample size varied according to follow-up availability. Participant selection and the analysis methodology are summarised in Figure 1.

**Figure 1.**
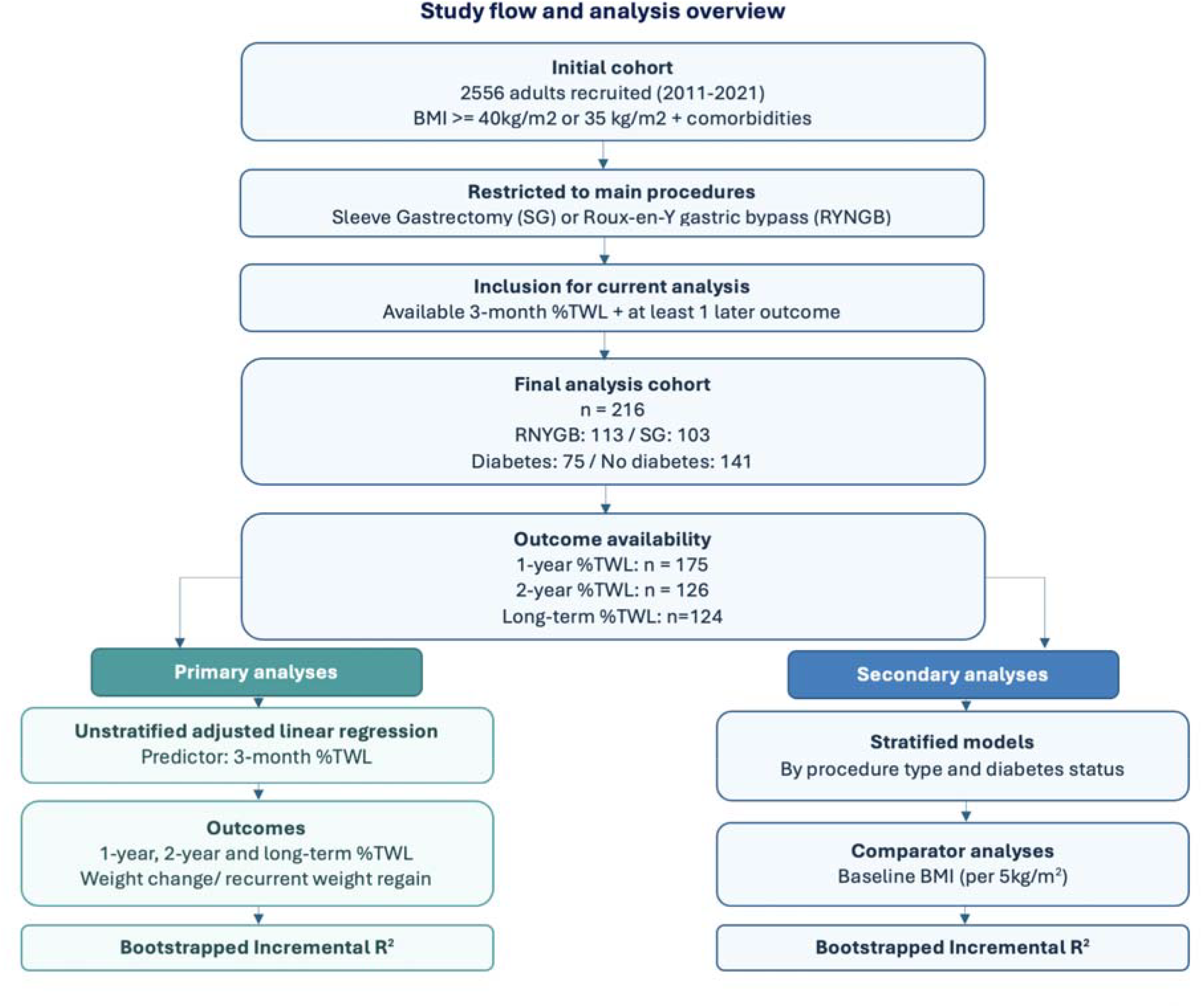
Study flow and analysis overview. Flowchart summarising participant selection from the initial cohort to the final analysis cohort, availability of 1-year, 2-year and long-term %TWL outcomes, and the primary and secondary analysis framework. Primary analyses assessed 3-month %TWL as a predictor of later %TWL and subsequent weight change/regain in the overall cohort. Secondary analysis included procedure- and diabetes-stratified models and baseline BMI comparator analyses. %TWL: percentage total weight loss, BMI: body mass index.

### Follow-up timepoints and weight definitions

Demographic, clinical, procedure and weight data were obtained from the study database and linked clinical records. Baseline weight was defined as a measurement obtained from 30 days before to 7 days after surgery. Postoperative measurements were grouped into 3-month, 1-year, 2-year and long-term (≥ 3 years) timepoints. Where several measurements were available, the value closest to the target timepoint was selected at 3-months, 1 year and 2 years; the most recent measurement was used for long-term follow-up. Mean long-term follow-up was 105.4 months (Range 35.8-160.2).

### Measured outcomes

The main weight-loss outcome was %TWL calculated as: %TWL = ((baseline weight-follow-up weight) / baseline weight) x 100.

Later weight-loss outcomes were 1-year %TWL, 2-year %TWL and long-term %TWL. Subsequent weight change/regain was calculated as the difference in %TWL between 1 and 2 years, 1 year and long-term follow-up, and 2-years and long-term follow-up. Negative values indicated recurrent weight gain, while positive values indicated continued weight loss.

### Predictors and covariates

The primary predictor was 3-month %TWL. Adjusted models included age at surgery, sex, procedure type, diabetes status, baseline BMI and ethnicity. Long-term models were additionally adjusted for months since surgery and change models for the number of months between the relevant measurements. These adjustments were selected to address clinical confounding and variation in follow-up timing.

Linear regression assessed associations between 3-month %TWL and each later outcome. Unadjusted and adjusted coefficients were reported with standard errors, 95% confidence intervals (CI) and p-values. Adjusted incremental R^2^ was calculated as the difference between R^2^ of the full model containing 3-month %TWL and covariates using 10,000 bootstrap resamples with paired bootstrap comparisons for selected incremental R^2^ estimates.

Secondary analyses repeated models after stratification by procedure type and diabetes status. Baseline BMI comparator models treated BMI per 5kg/m2 as the exposure and used the corresponding adjustment set without baseline BMI as the covariate. Full methods and results for these analyses are provided in the Supplementary Material.

Analyses were conducted in R^[10]^, with data management using tidyverse packages^[11]^ and visualisation using ggplot2^[12]^. Statistical significance was defined as p < 0.05.

## Results

### Association between 3-month %TWL and later %TWL

Greater 3-month %TWL was associated with higher 1-year and 2-year %TWL. After adjustment, each 1-percentage-point increase in 3-month %TWL was associated with a 0.95-percentage-point higher 1-year %TWL (SE = 0.08, 95% CI 0.78 to 1.08, p = 2.005e-24). 0.59 percentage-points higher 2-year %TWL in the adjusted model (SE = 0.13, 95% CI 0.33 to 0.85, p = 2.023e-05). The association between 3-month %TWL and longer-term %TWL was not statistically significant (β = 0.32, SE = 0.18, 95% CI -0.04 to 0.67, p = 0.078).

Adjusted R^2^ analysis was 38.23% for 1-year %TWL, 11.75% for 2-year %TWL and 2.25% for longer-term %TWL. The paired bootstrap comparison showed a significant difference between the 1-year and 2-year estimates, indicating that the additional explanatory value of 3-month %TWL was greatest at nearer follow-up. (Figure 2A).

**Figure 2.**
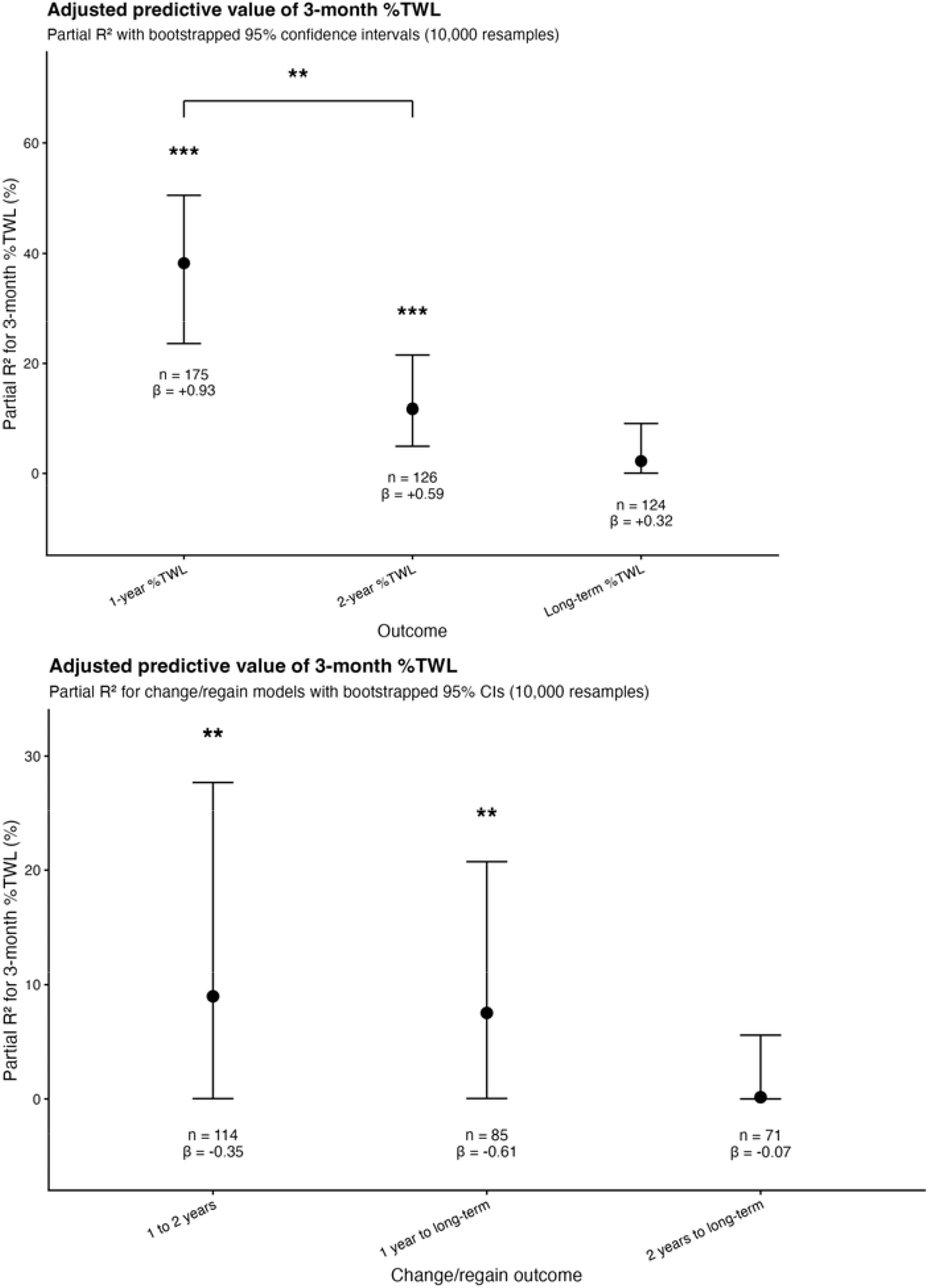
A-B. Adjusted incremental R^2^ for 3-month %TWL predicted later weight loss and subsequent weight change/regain outcomes. **(A)**Adjusted incremental R^2^ for 3-month %TWL predicting later %TWL at 1-year, 2-year, and long-term follow-up. Bracket indicates a statistically significant difference in incremental R^2^ between 1-year and 2-year %TWL outcomes **(B)**Adjusted incremental R^2^ for 3-month %TWL predicting subsequent weight change/regain outcomes. Points show the additional variance explained by the 3-month %TWL Later %TWL models were adjusted for age at surgery, sex, procedure type, diabetes status, baseline BMI and ethnicity, with long-term %TWL models being additionally adjusted for months since surgery. Change/regain models were additionally adjusted for the interval between the relevant follow-up measurements. Error bars show bootstrapped 95% CI. β values displayed below each point represent the adjusted regression coefficient for 3-month %TWL Significance stars indicate the p-value for the adjusted β-coefficient. *** = p < 0.001, ** = p < 0.01. For change/regain outcomes, negative β indicates greater weight regain. Model-adjusted mean ± standard error (SE) for each procedure-stratified outcome is reported in Supplementary Table 1.

### Association with subsequent recurrent weight change/gain

Greater 3-month %TWL was associated with greater subsequent recurrent weight gain from 1-year to long-term follow-up. With each 1 percentage point increase in 3-month %TWL with 0.35 percentage point lower change from 1 to 2 years (SE = 0.10, 95% CI -0.55 to -0.14, p = 0.00126299) and a 0.61 percentage point lower change in %TWL from 1-year to long-term (SE = 0.23, 95% CI -1.07 to -0.16, p = 0.009103). There was no clear association with change from 2 years to long-term follow-up (β = - 0.07, SE = 0.21, 95% CI -0.49 to 0.35, p = 0.72759996).

Adjusted incremental R^2^ was 8.98% for change from 1 to 2 years, 7.51% for change from 1 year to long-term follow-up and 0.15% for change from 2 years to long-term follow-up. Bootstrapped CIs overlapped, providing no clear evidence that the additional variance explained by 3-month %TWL differed across intervals (Figure 2B).

### Secondary analyses

Procedure- and diabetes-stratified analyses were broadly consistent with the primary analyses’ findings: 3-month %TWL predicted 1-year and 2-year %TWL in both procedure and diabetes groups, while subgroup estimates were less precise and there were no long-term associations. Baseline BMI was significantly associated with 3-month %TWL and with 1-year %TWL in some models, but 3-month %TWL explained substantially more additional variance in 1-year %TWL. Full estimates, bootstrap comparisons and figures are presented in Supplementary Figures S1-S6 and Supplementary Tables S1-S3.

## Discussion

### Principal findings

In this cohort, 3-month %TWL was strongly associated with 1-year and 2-year %TWL but not with long-term %TWL. Its adjusted incremental R^2^ was greatest for 1-year outcomes and declined at later follow-up. Greater early weight loss was also associated with greater recurrent weight gain from 1 to 2 years and from 1 year to long-term follow-up. Together, these findings indicate that 3-month %TWL is informative about short- to medium-term weight trajectories, while its association with weight outcomes attenuates over longer-term follow-up.

### Prediction of later weight-loss outcomes

The association with 1-year and 2-year outcomes is consistent with previous evidence that early postoperative weight loss predicts later MBS outcomes^[5,13]^. Longer-term evidence is less consistent: Tettero et al. found that 3-month %TWL remained associated with weight-loss trajectories up to 5 years after RYNGB and SG^[14]^, whereas another study found no association between early weight loss at 4 weeks and 7-year outcomes after SG^[15]^. Differences may reflect variation in the timing of early weight assessment, procedure type and duration of follow-up. In the present study, the greater incremental R^2^ at 1 year suggests that 3-month %TWL may reflect the initial response to surgery, whereas longer-term outcomes reflect maintenance behaviours, engagement with follow-up, psychological factors and subsequent management strategies^[16,17]^. Postoperative weight trajectories may also become increasingly heterogeneous over time, with distinct patterns of weight loss and regain previously described^[18]^.

### Early weight loss and subsequent recurrent weight gain

Greater 3-month %TWL was further associated with greater recurrent weight gain over some intervals. This does not mean that patients with greater early weight loss had poorer absolute outcomes, but rather that those who lost more weight initially may have had greater scope for measurable regain after approaching nadir weight. Previous research has similarly linked greater early postoperative weight loss with both lower nadir weight and later weight regain^[19]^, while another study reported that patients with lower 3-month %TWL had poorer absolute weight-loss outcomes but greater subsequent weight stability^[14]^. Together, these findings suggest that the magnitude of early weight loss and then the following weight stability may represent related but different aspects of postoperative weight trajectory. Therefore, a strong early weight-loss response should not be interpreted as removing the need for continued long-term monitoring.

### Secondary and comparator analyses

Secondary analyses suggested that the main short- and medium-term associations were present after stratification by procedure type and diabetes status, although smaller subgroup samples produced wider CIs. These findings should be interpreted with caution until formal evidence shows that subgroup effects differ. The baseline BMI analyses also showed that early %TWL was not entirely independent of starting BMI. Nevertheless, 3-month %TWL explained substantially more additional variance in 1-year %TWL than baseline BMI, supporting its value as a postoperative marker instead of simply reflecting starting BMI^[8,9]^. Full secondary analyses can be found in the Supplementary Material.

### Strengths and limitations

Strengths of this work include assessment of a routinely recorded measure across fixed 1-year, 2-year and long-term outcomes, inclusion of subsequent weight change outcomes, multivariable adjustment for clinically relevant factors and bootstrapped R^2^ estimates. The study also included both SG and RNYGB, within a multi-ethnic UK cohort.

Limitations include a modest sample size, particularly limiting for subgroup and long-term analyses, and outcome-specific loss to follow-up. Long-term results may be affected by selection bias if patients with available measurements differed from those without follow-up, and adjustment for follow-up timing does not remove bias caused by missing outcomes. Residual confounding is also possible, including from behavioural, psychological and postoperative treatment factors not consistently available. Since the cohort was recruited from NHS bariatric services, the findings may be most applicable to similar settings, and further work should be done in other populations.

### Clinical implications and future research

Clinically, 3-month %TWL may be an early indicator of later weight trajectories. Patients with lower 3-month %TWL could be considered for earlier review or additional support, while those with a strong early response might still require long-term monitoring for recurrent weight gain. Human studies have demonstrated inter-individual differences in weight variability, with greater variability associated with subsequent weight gain and poorer long-term weight maintenance^[20,21]^. Heterogeneity in postoperative weight trajectories has also been reported following MBS^[18]^. Experimental work in mice has similarly demonstrated variation in weight-stability and recovery traits influenced by genetic and environmental factors^[22]^, although the relevance of these findings to postoperative weight trajectories in humans remains uncertain. Therefore, further research should investigate the factors underlying this variability and whether early postoperative weight-loss patterns can identify patients who would benefit from more intensive long-term management.

## Conclusion

Three-month %TWL predicted 1-year and 2-year weight-loss outcomes after MBS, although its association with %TWL was not maintained beyond 3 years. It explained more additional variance in 1-year %TWL than baseline BMI, supporting its use as an early postoperative marker. Its association with recurrent weight gain, together with recognised heterogeneity in postoperative weight stability, highlights the importance of continued long-term follow-up and warrants further investigation into whether early weight-loss patterns can identify patients requiring more intensive long-term weight management.

## Data Availability

Data availability for this study is subject to ethical restrictions under the Personalised Medicine for Morbid Obesity study (ClinicalTrials.gov: NCT01365416). Researchers wishing to access the data may apply by contacting Prof. Alexandra Blakemore at. Access is subject to appropriate ethical and governance approvals.

## Declarations

## Ethical approval

Ethical approval was granted by the NRES Committee for London Riverside (REC reference 11/LO/0935), and the research was conducted in accordance with the Declaration of Helsinki.

## Trial registration

https://clinicaltrials.gov/study/NCT01365416

## Informed consent

Informed consent was obtained from all participants recruited to the Personalised Medicine for Morbid Obesity cohort.

## Data availability

The data supporting the findings of this study are not publicly available due to participant confidentiality and ethical restrictions. Data may be available from the corresponding author upon reasonable request and subject to appropriate approvals

