## Supplementary Information for "Greater early weight loss after metabolic/bariatric surgery predicts greater loss up to 2 years but is also associated with weight regain in the longer term"

**Supplementary Material**

Supplementary methods

Procedure-stratified analyses were performed separately in patients undergoing sleeve gastrectomy and Roux-en-Y gastric bypass. Diabetes-stratified analyses were also performed separately in patients with and without diabetes. Baseline BMI comparator models were fitted using baseline BMI per 5kg/m2 as the main predictor. These models used the same adjustment approach as the main models, except the stratification variable was removed from the relevant stratified model and baseline BMI was not included. Incremental R^2^ was calculated as the difference in R^2^ between the full adjusted was calculated as the difference in R^2^ between the full adjusted model and the corresponding covariate-only model, with 95% confidence intervals estimated using 10,000 bootstrap resamples.

**Figure S1A-B. Procedure-stratified adjusted incremental R^2^ for 3-month %TWL predicting later weight-loss and subsequent weight change/regain outcomes**

**(A)** Adjusted incremental R^2^ for 3-month %TWL predicted later %TWL at 1-year, 2-year and long-term follow-up, stratified by procedure type

**(B)** Adjusted incremental R^2^ for 3-month %TWL predicting subsequent weight change/regain outcomes, stratified by procedure type

Points show additional variance explained by 3-month %TWL within procedure-stratified models. Later %TWL models were adjusted for age at surgery, sex, diabetes status, baseline BMI and ethnicity, with long-term %TWL models being additionally adjusted for months since surgery. Change/regain models were additionally adjusted for the interval between the relevant follow-up measurements.

Error bars show bootstrapped 95% CI. Β values displayed below each point represent the adjusted regression coefficient for 3-month %TWL.

Significance stars indicate the p-value for the adjusted β-coefficient. *** = p < 0.001, ** = p < 0.01, * = p < 0.05.

For change/regain outcomes, negative β indicates greater weight regain.

Model-adjusted mean ± standard error (SE) for each procedure-stratified outcome is reported in Supplementary Table 2.

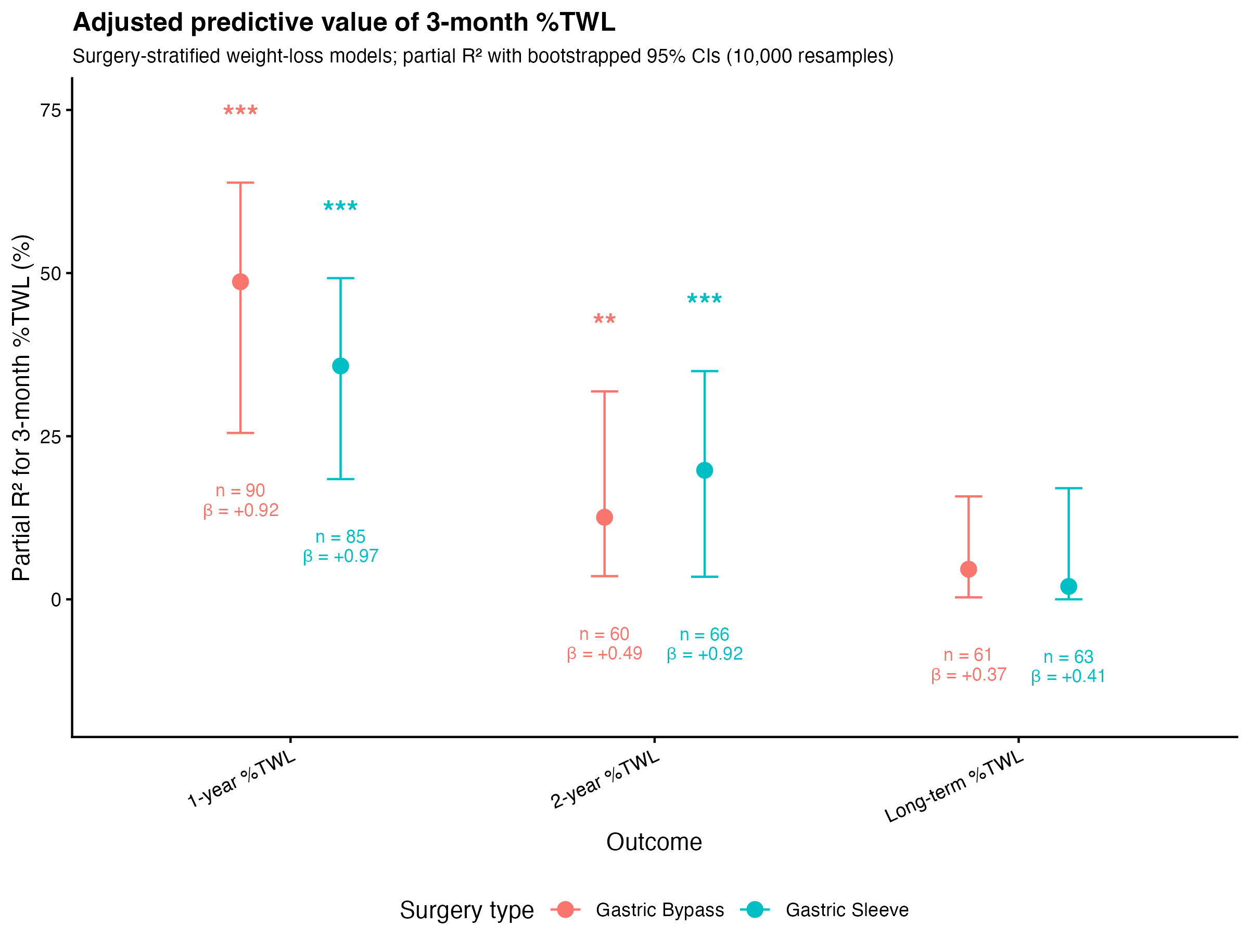

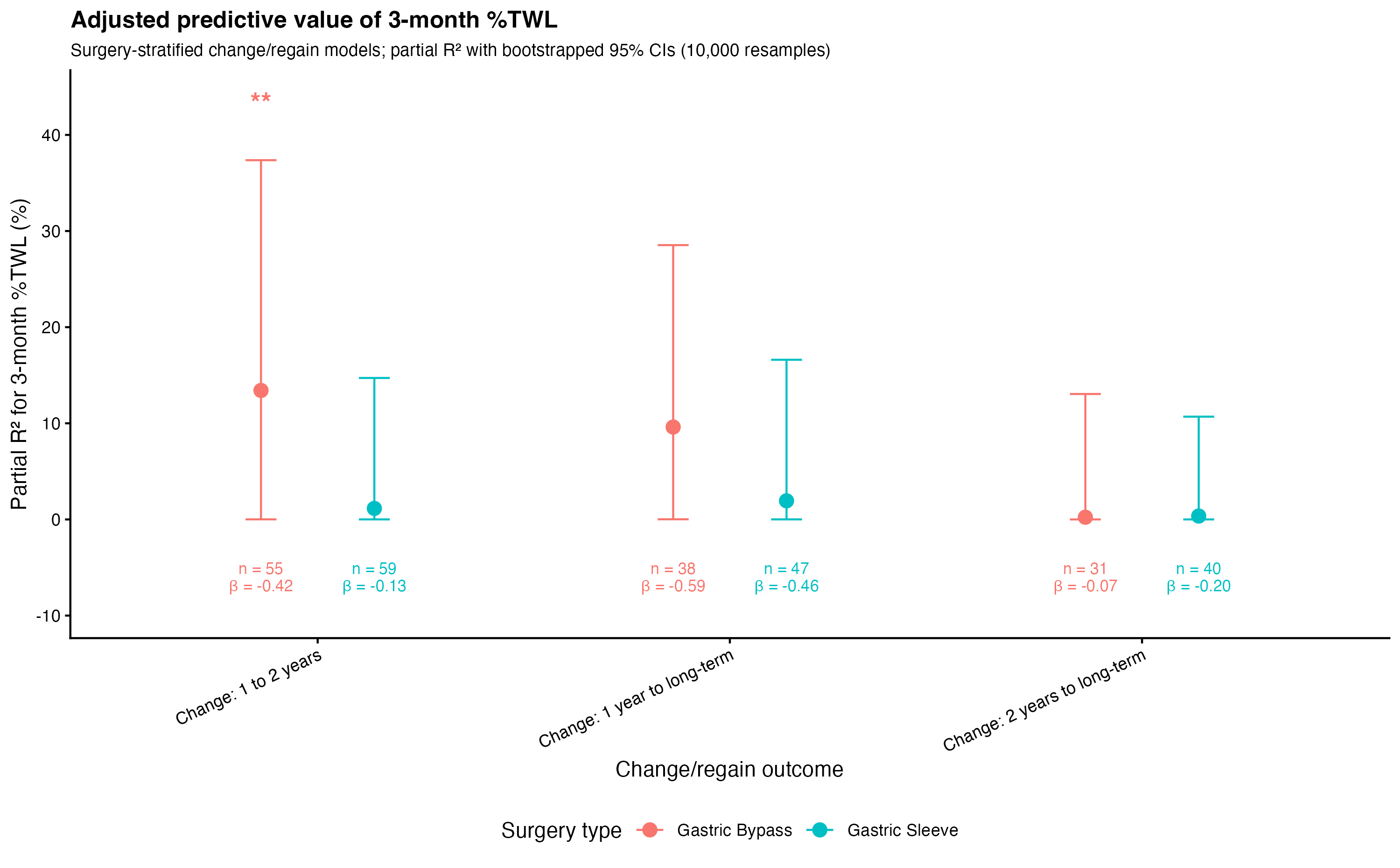

**Figure S2A-B. Diabetes-stratified adjusted incremental R^2^ for 3-month %TWL predicting later weight-loss and subsequent weight change/regain outcomes**

**(A)** Adjusted incremental R^2^ for 3-month %TWL predicted later %TWL at 1-year, 2-year and long-term follow-up, stratified by diabetes status

**(B)** Adjusted incremental R^2^ for 3-month %TWL predicting subsequent weight change/regain outcomes, stratified by diabetes status

Points show additional variance explained by 3-month %TWL within procedure-stratified models. Later %TWL models were adjusted for age at surgery, sex, procedure type, baseline BMI and ethnicity, with long-term %TWL models being additionally adjusted for months since surgery. Change/regain models were additionally adjusted for the interval between the relevant follow-up measurements.

Error bars show bootstrapped 95% CI. Β values displayed below each point represent the adjusted regression coefficient for the 3-month %TWL

Significance stars indicate the p-value for the adjusted β-coefficient. *** = p < 0.001, ** = p < 0.01, * = p < 0.05.

Model-adjusted mean ± standard error (SE) for each procedure-stratified outcome is reported in Supplementary Table 3.

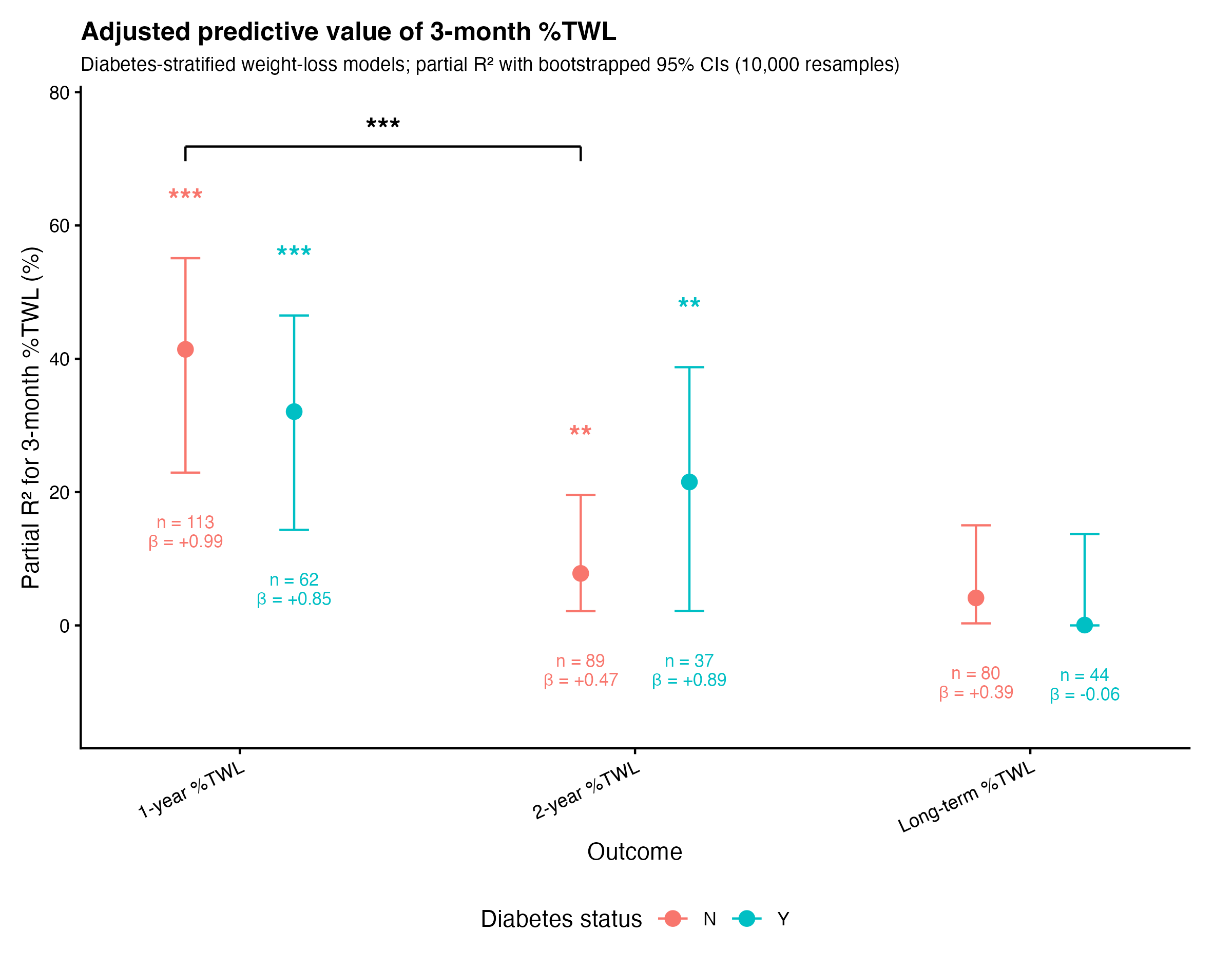

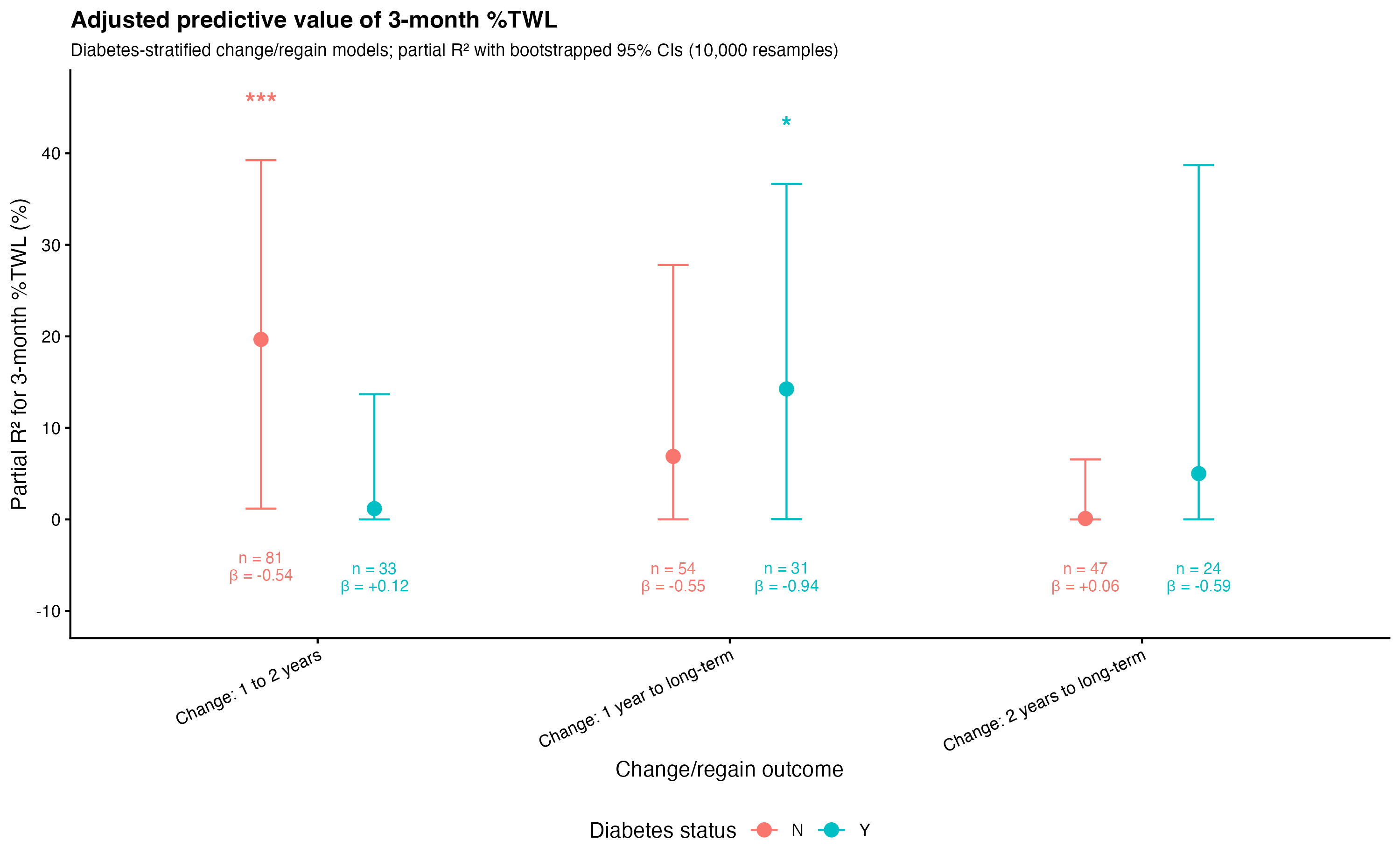

**Figure S3. Adjusted association between baseline BMI and 3-month %TWL.**

The point shows the adjusted regression coefficient for baseline BMI modelled per 5kg/m2 increase. Error bars show 95% confidence intervals. The dashed horizontal line represents no association. Values above zero indicate that higher baseline BMI was associated with greater 3-month %TWL. The model was adjusted for age at surgery, sex, procedure type, diabetes status and ethnicity.

*** = p < 0.001

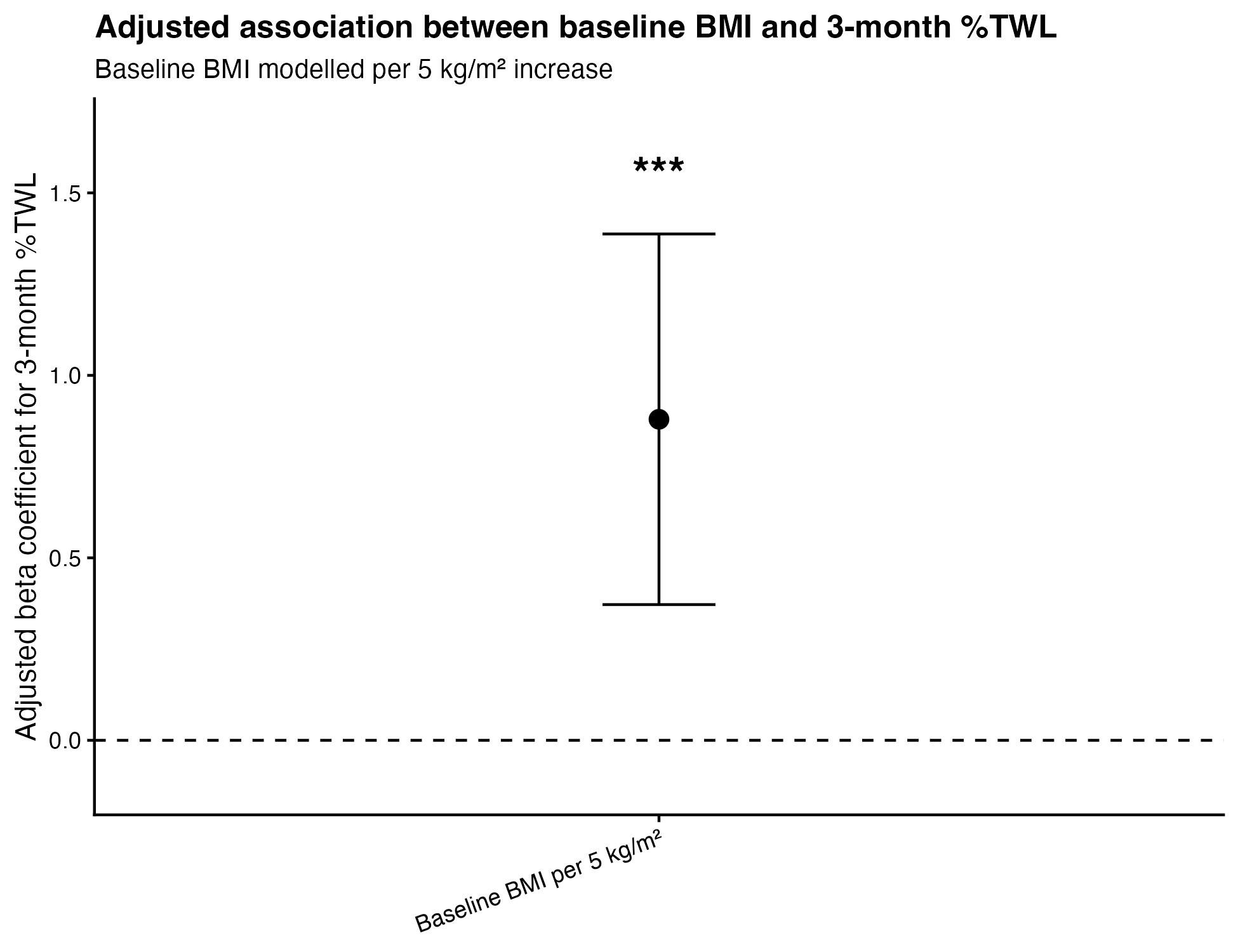

**Figure S4A-B. Baseline BMI as a comparator predictor of later weight loss and subsequent weight change/regain outcomes.**

(**A**)Adjusted incremental R^2^ for baseline BMI predicting later %TWL at 1 year, 2 years and long-term follow-up.

(**B**) Adjusted incremental R^2^ for baseline BMI predicting subsequent weight change/regain outcomes

Points show additional variance explained by baseline BMI. Later %TWL models were adjusted for age at surgery, sex, procedure type, diabetes status, and ethnicity, with long-term %TWL models being additionally adjusted for months since surgery. Change/regain models were additionally adjusted for the interval between the relevant follow-up measurements.

Error bars show bootstrapped 95% CI. β values displayed below each point represent the adjusted regression coefficient for 3-month %TWL

Significance stars indicate the p-value for the adjusted β-coefficient. *** = p < 0.001, ** = p < 0.01.

For change/regain outcomes, a negative β indicates greater weight regain.

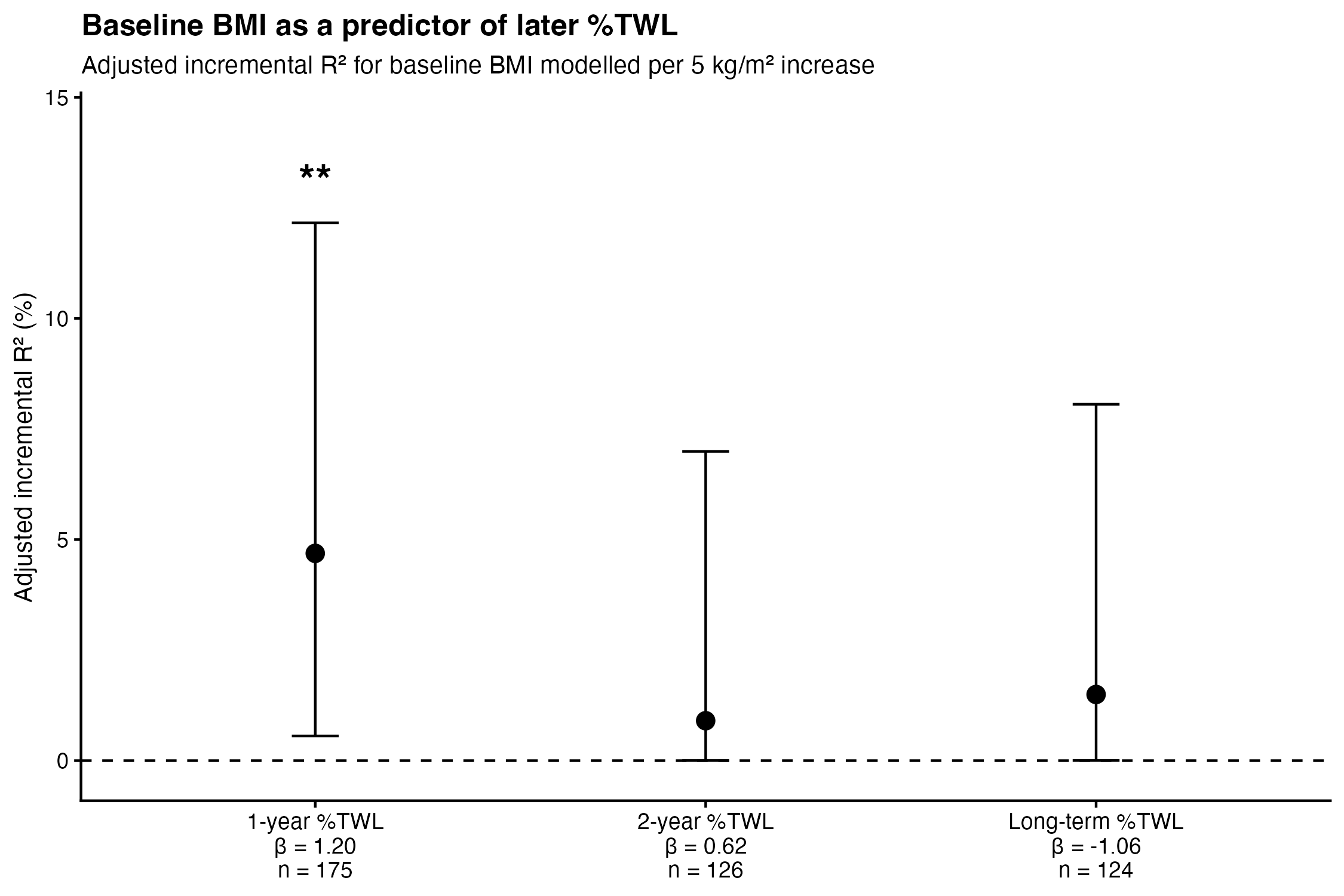

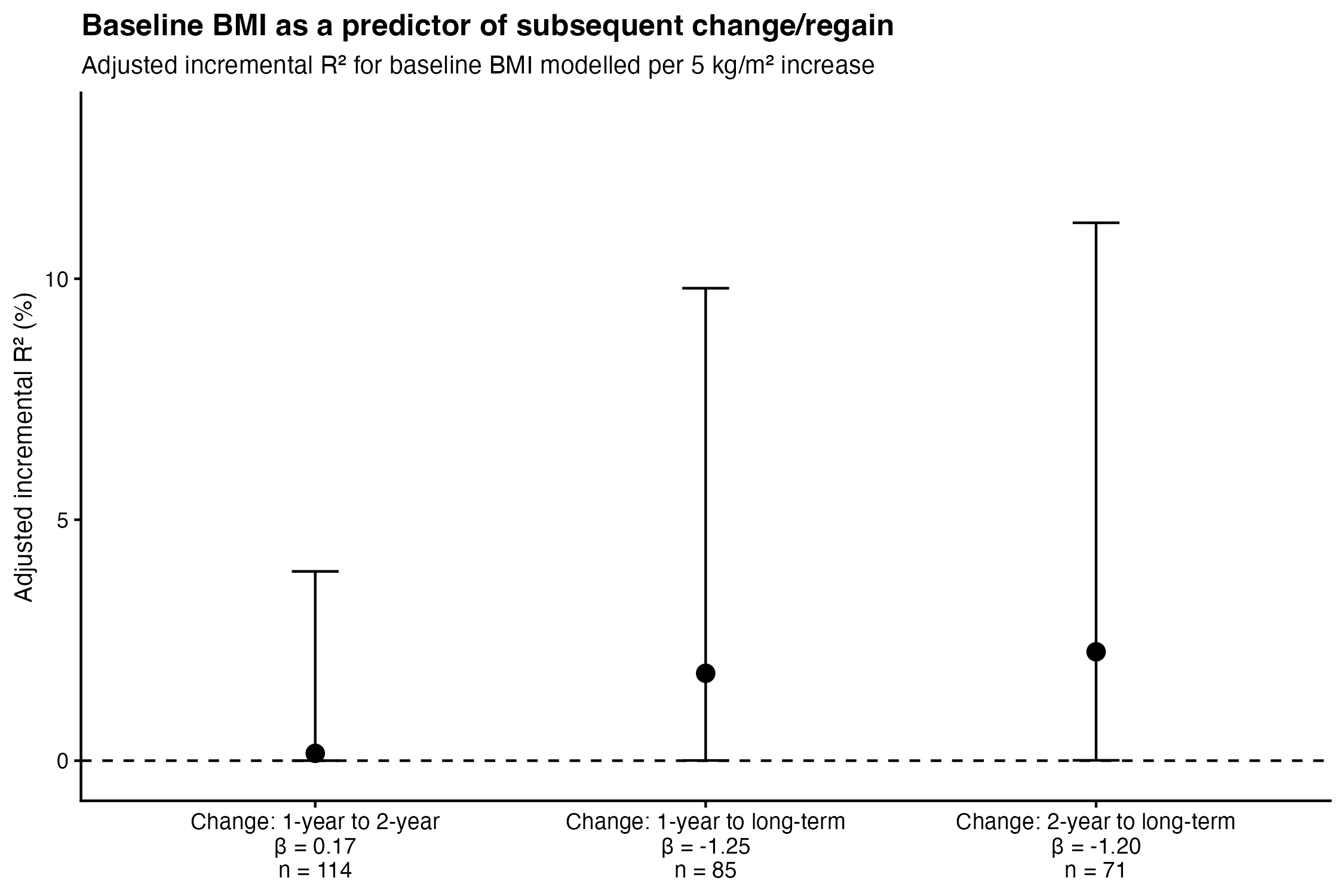

**Supplementary Figure S5A-B. Procedure-stratified baseline BMI comparator models for later weight loss and subsequent change/regain outcomes**

(**A**)Adjusted incremental R^2^ for baseline BMI predicting later %TWL at 1-year, 2-year and long-term follow-up, stratified by procedure type

(**B**) Adjusted incremental R^2^ for baseline BMI predicting subsequent change/regain outcomes, stratified by procedure type

Points show additional variance explained by baseline BMI. Later %TWL models were adjusted for age at surgery, sex, diabetes status, and ethnicity, with long-term %TWL models being additionally adjusted for months since surgery. Change/regain models were additionally adjusted for the interval between the relevant follow-up measurements.

Error bars show bootstrapped 95% CI. β values displayed below each point represent the adjusted regression coefficient for 3-month %TWL.

Significance stars indicate the p-value for the adjusted β-coefficient. * = p < 0.05.

For change/regain outcomes, negative β indicates greater weight regain.

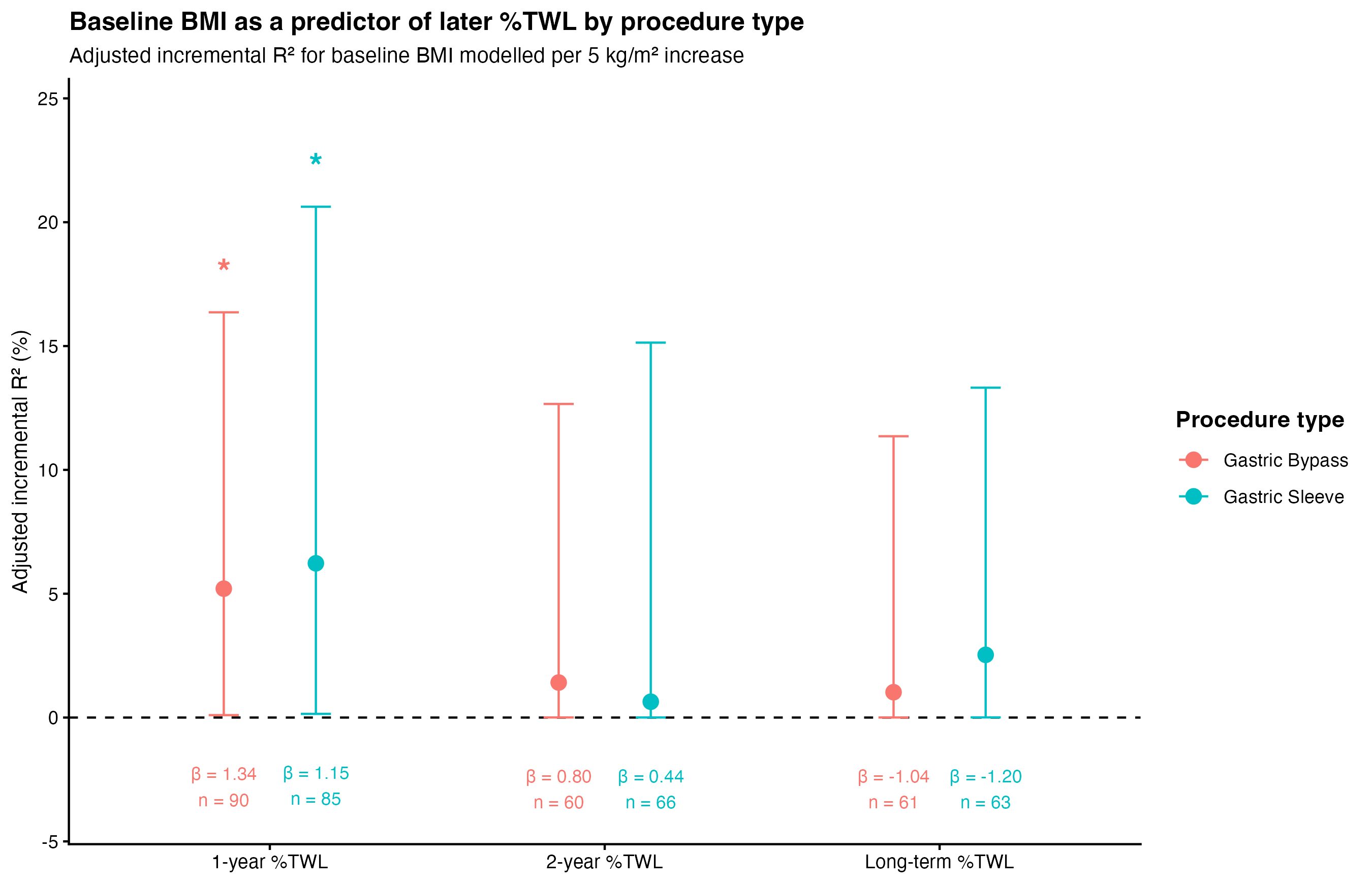

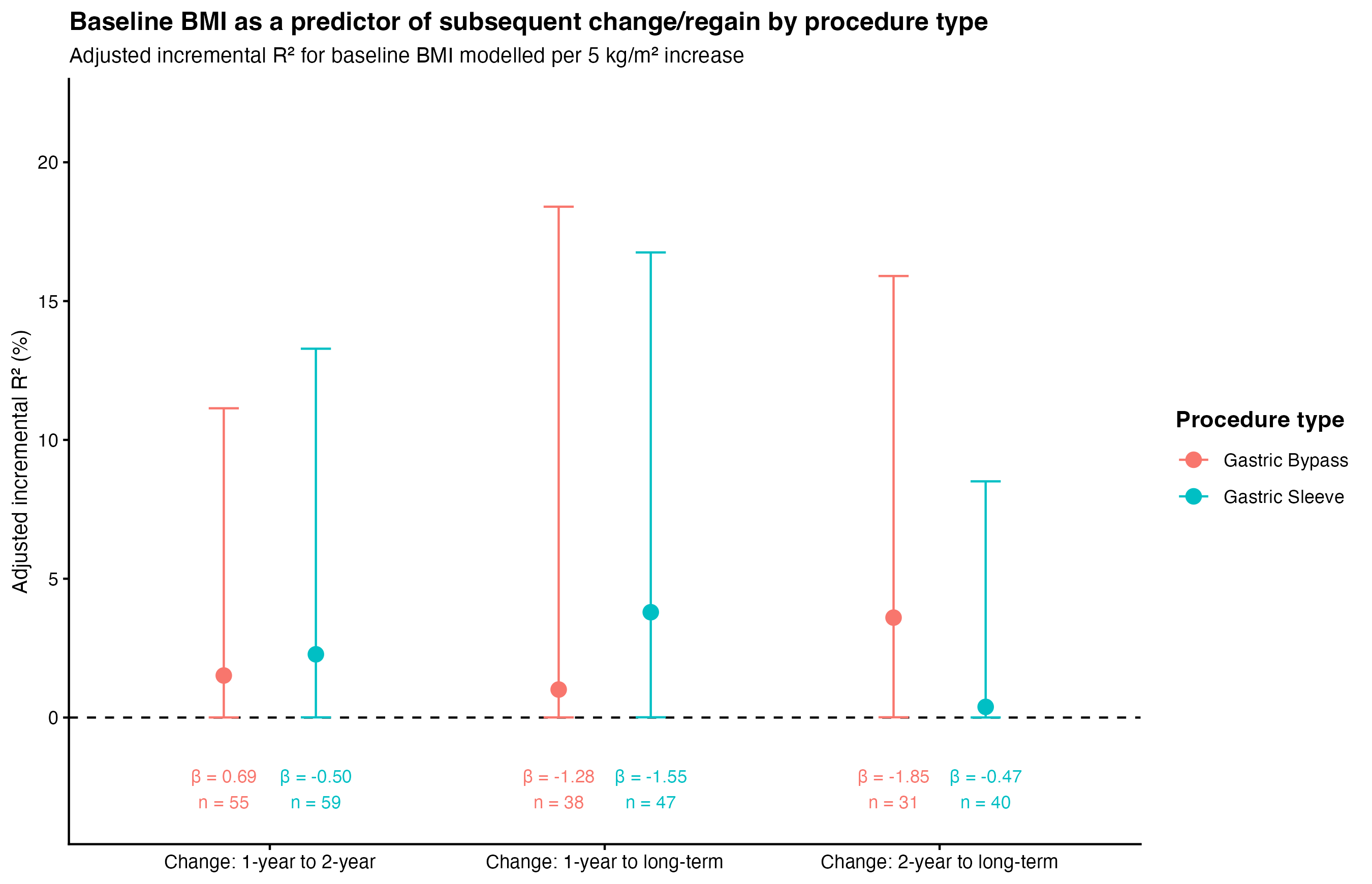

**Supplementary Figure S6A-B. Diabetes-stratified baseline BMI comparator models for later weight loss and subsequent change/regain outcomes**

(**A**)Adjusted incremental R^2^ for baseline BMI predicting later %TWL at 1-year, 2-year and long-term follow-up, stratified by procedure type

(**B**) Adjusted incremental R^2^ for baseline BMI predicting subsequent change/regain outcomes, stratified by procedure type

Points show additional variance explained by baseline BMI. Later %TWL models were adjusted for age at surgery, sex, procedure type, ethnicity, with long-term %TWL models being additionally adjusted for months since surgery. Change/regain models were additionally adjusted for the interval between the relevant follow-up measurements.

Error bars show bootstrapped 95% CI. β values displayed below each point represent the adjusted regression coefficient for 3-month %TWL.

Significance stars indicate the p-value for the adjusted β-coefficient. ** = p < 0.01, * = p < 0.05.

For change/regain outcomes, negative β indicates greater weight regain.

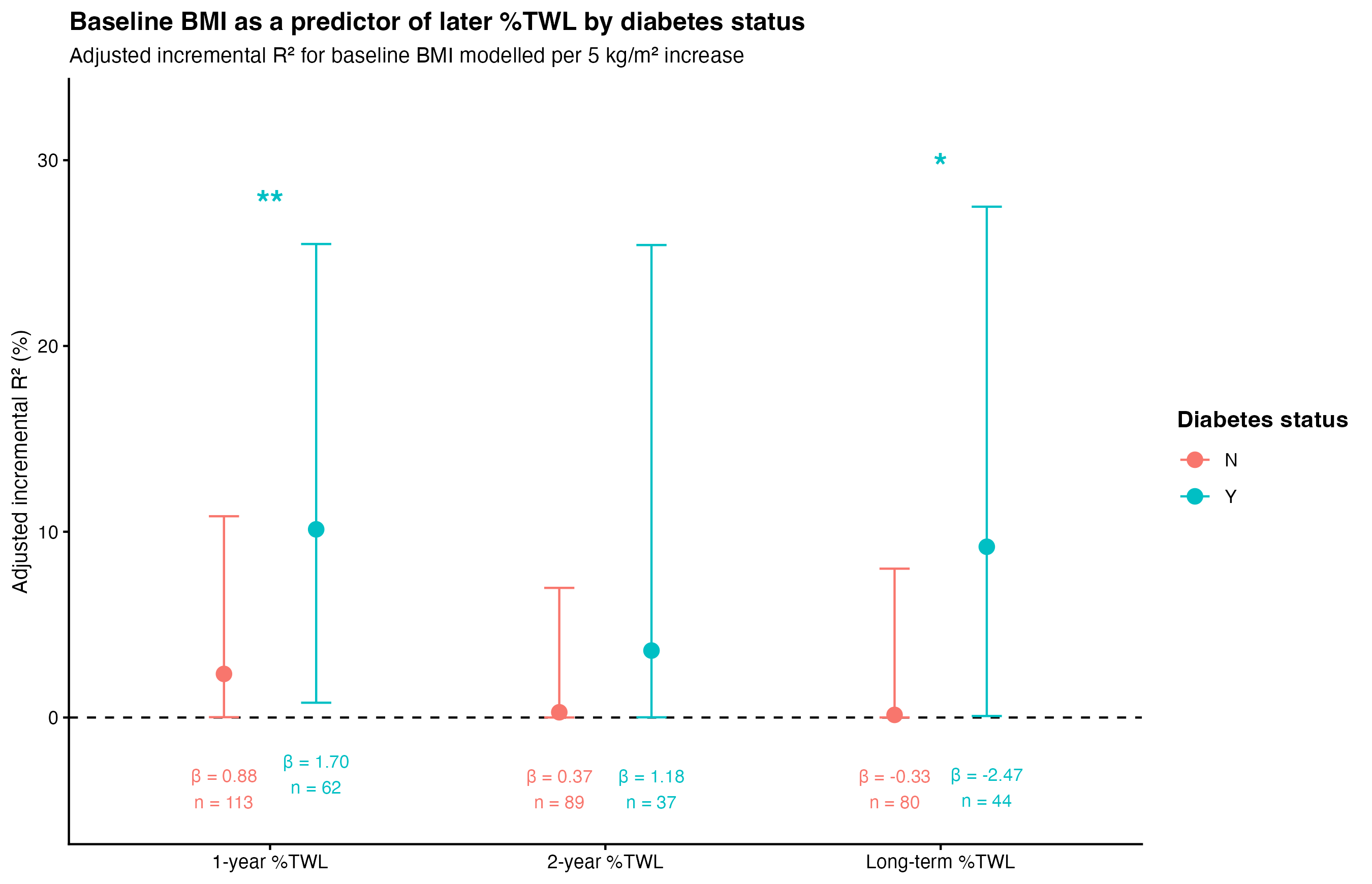

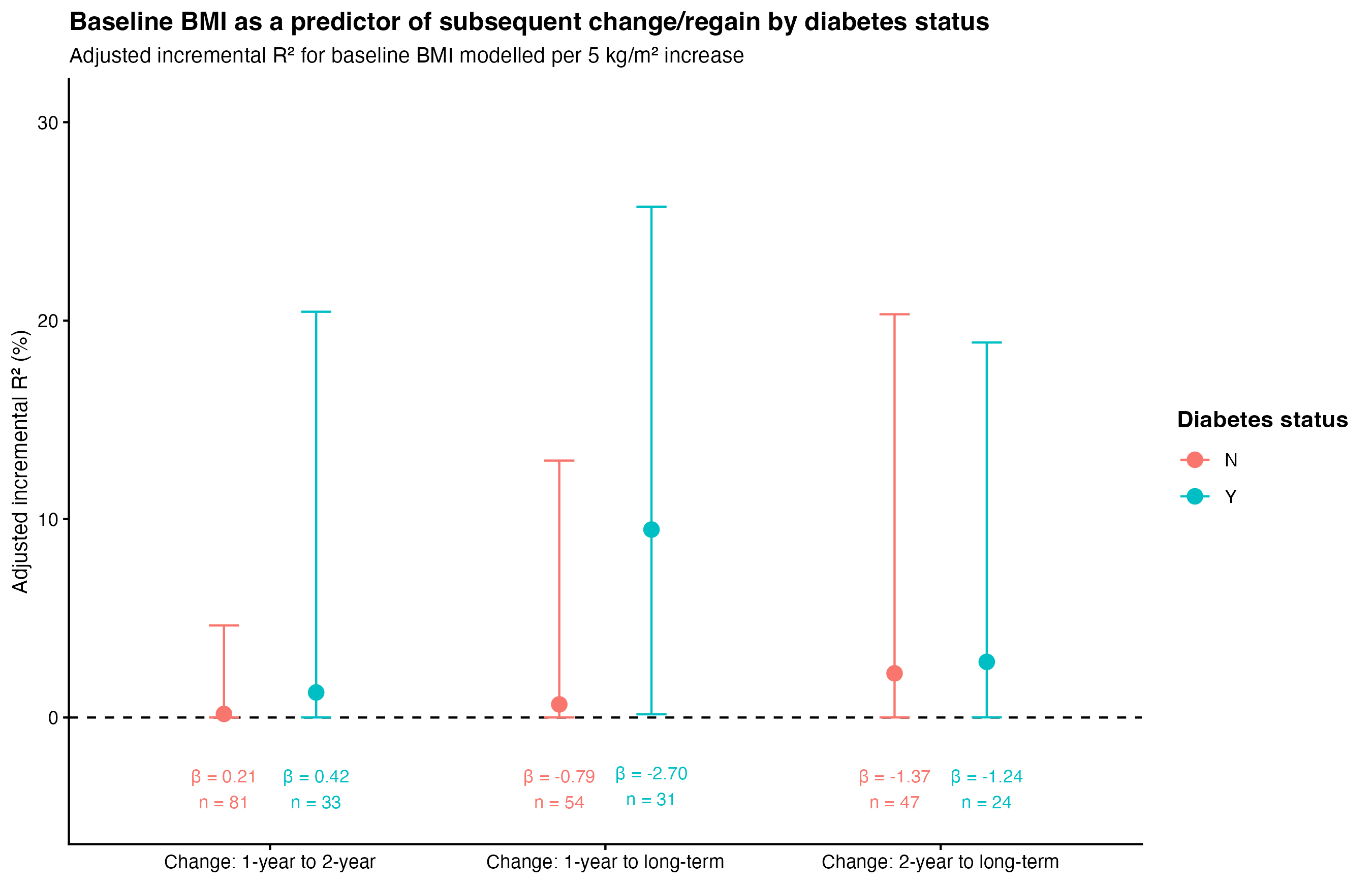

**Supplementary Table 1. Model adjusted mean outcomes and standard errors for unstratified weight loss and change/regain outcomes.**

Model-adjusted mean ± standard error (SE) values are shown for the unstratified outcomes presented in Figures 1A and 1B. Adjusted means were estimated from the corresponding adjusted linear regression models using the same covariate adjustment described in Figure 1. For change/regain outcomes, negative values indicate weight regain, while positive values indicate continued weight loss.

| **Outcome** | **Adjusted mean ± SE** | **95% CI** | **Timing adjustment** |
| --- | --- | --- | --- |
| **1-year %TWL** | 28.37 ± 0.43 | 27.52 to 29.21 | No additional timing covariate |
| **2-year %TWL** | 27.21 ± 0.75 | 25.73 to 28.68 | No additional timing covariate |
| **Long-term %TWL** | 20.87 ± 1.02 | 18.85 to 22.89 | Months since surgery long-term |
| **Change: 1-year to 2-year** | -0.61 ± 0.60 | -1.80 to 0.59 | Months between 1y and 2y |
| **Change: 1-year to Long-term** | -6.64 ± 1.30 | -9.24 to -4.05 | Months between 1y and long-term |
| **Change: 2-years to long-term** | -4.64 ± 1.22 | -7.08 to -2.19 | Months between 2y and long-term |

**Supplementary Table 2. Model-adjusted mean outcomes and standard errors for procedure-stratified weight-loss and change/regain outcomes.**

Model-adjusted mean ± standard error (SE) values are shown for procedure-stratified outcomes in Figures S1A and S1B. Adjusted means were estimated from the corresponding procedure-stratified adjusted linear regression models using the same covariates as described for Figure 2. For change/regain outcomes, negative values indicate weight regain, while positive values indicate continued weight loss.

| **Outcome** | **Procedure type** | **Adjusted mean ± SE** | **95% CI** | **Timing adjustment** |
| --- | --- | --- | --- | --- |
| **1-year %TWL** | Gastric Bypass | 31.12 ± 0.61 | 29.91 to 32.34 | No additional timing covariate |
| **1-year %TWL** | Gastric Sleeve | 25.45 ± 0.61 | 24.24 to 26.66 | No additional timing covariate |
| **2-year %TWL** | Gastric Bypass | 31.26 ± 1.14 | 28.97 to 33.54 | No additional timing covariate |
| **2-year %TWL** | Gastric Sleeve | 23.53 ± 0.98 | 21.56 to 25.49 | No additional timing covariate |
| **Long-term %TWL** | Gastric Bypass | 23.51 ± 1.44 | 20.64 to 26.39 | Months since surgery long-term |
| **Long-term %TWL** | Gastric Sleeve | 18.31 ± 1.42 | 15.47 to 21.15 | Months since surgery long-term |
| **Change: 1-year to 2-year** | Gastric Bypass | 0.60 ± 0.98 | -1.38 to 2.57 | Months between 1y and 2y |
| **Change: 1-year to 2-year** | Gastric Sleeve | -1.73 ± 0.68 | -3.08 to -0.37 | Months between 1y and 2y |
| **Change: 1-year to long-term** | Gastric Bypass | -6.00 ± 2.07 | -10.22 to -1.77 | Months between 1y and long-term |
| **Change: 1-year to long-term** | Gastric Sleeve | -7.17 ± 1.62 | -10.45 to -3.88 | Months between 1y and long-term |
| **Change: 2-year to long-term** | Gastric Bypass | -6.32 ± 1.96 | -10.37 to -2.28 | Months between 2y and long-term |
| **Change: 2-year to long-term** | Gastric Sleeve | -3.33 ± 1.64 | -6.67 to 0.01 | Months between 2y and long-term |

**Supplementary Table 3. Model-adjusted mean outcomes and standard errors for diabetes-stratified weight-loss and change/regain outcomes.**

Model-adjusted mean ± standard error (SE) values are shown for procedure-stratified outcomes in Figures S2A and S2B. Adjusted means were estimated from the corresponding diabetes-stratified adjusted linear regression models using the same covariates as described for Figure 3. For change/regain outcomes, negative values indicate weight regain, while positive values indicate continued weight loss.

| **Outcome** | **Diabetes status** | **Adjusted mean ± SE** | **95% CI** | **Timing adjustment** |
| --- | --- | --- | --- | --- |
| **1-year %TWL** | Y | 28.65 ± 0.74 | 27.17 to 30.13 | No additional timing covariate |
| **1-year %TWL** | N | 28.21 ± 0.52 | 27.18 to 29.25 | No additional timing covariate |
| **2-year %TWL** | Y | 28.23 ± 1.28 | 25.62 to 30.85 | No additional timing covariate |
| **2-year %TWL** | N | 26.78 ± 0.90 | 24.98 to 28.58 | No additional timing covariate |
| **Long-term %TWL** | Y | 25.33 ± 1.56 | 22.17 to 28.49 | Months since surgery long-term |
| **Long-term %TWL** | N | 18.42 ± 1.32 | 15.79 to 21.05 | Months since surgery long-term |
| **Change: 1-year to 2-year** | Y | 0.04 ± 1.03 | -2.08 to 2.16 | Months between 1y and 2y |
| **Change: 1-year to 2-year** | N | -0.87 ± 0.70 | -2.26 to 0.52 | Months between 1y and 2y |
| **Change: 1-year to long-term** | Y | -2.51 ± 1.88 | -6.41 to 1.38 | Months between 1y and long-term |
| **Change: 1-year to long-term** | N | -9.02 ± 1.77 | -12.57 to -5.46 | Months between 1y and long-term |
| **Change: 2-year to long-term** | Y | -1.08 ± 2.08 | -5.49 to 3.33 | Months between 2y and long-term |
| **Change: 2-year to long-term** | N | -6.45 ± 1.59 | -9.68 to -3.23 | Months between 2y and long-term |
